# Deep sequencing reveals subpopulation dynamics associated with treatment failure in a rare non-tuberculous mycobacterial infection

**DOI:** 10.64898/2026.08.10.26359948

**Authors:** Aruna R. Menon, Carla Mariner-Llicer, Ana María Xet-Mull, Naseem Alavian, Mariana G. López, Eileen K. Maziarz, Mark J. Lee, David M. Tobin, Jason E. Stout, Iñaki Comas

**Affiliations:** Department of Molecular Genetics and Microbiology, Duke University School of Medicine, Durham, NC 27710, USA; Universitat de València, València, Spain; Institut de Biologia de Sistemes (I2SysBio), València, Spain; Mycobacterial Infections Study Group (GEIM) of the Spanish Society of Infectious Diseases and Clinical Microbiology (SEIMC), Spain; Division of Infectious Diseases and International Health, Department of Medicine, Duke University School of Medicine, Durham, NC 27710, USA; Instituto de Biomedicina de València (CSIC), València, Spain; Department of Pathology, Duke University School of Medicine, Durham, NC 27710, USA; Department of Integrative Immunobiology, Duke University School of Medicine, Durham, NC 27710, USA; CIBER in Epidemiology and Public Health, Madrid, Spain

**Keywords:** Nontuberculous mycobacteria, deep-sequencing, within-patient evolution, population dynamics, treatment failure, longitudinal analysis, drug-resistance

## Abstract

**Background:** Nontuberculous mycobacteria (NTM) are an increasingly common group of pathogens that remain challenging to diagnose and treat effectively. The lack of standardization of NTM management, from identification to antibiotic resistance prediction, results in imperfect correlations between treatment and outcomes. This study characterizes the genetic heterogeneity of a previously uncharacterized NTM during a 29-month bacteremia with acquired drug resistance.

**Results:** In contrast to the initial diagnostic result identifying *M. nebraskense*, a rare NTM causing disease in humans, whole genome sequencing (WGS) identified *Mycobacterium sp. SMC-2*, a species with only one publicly available genome. High-resolution analysis of variants revealed 444 unique SNPs and 26 indels in 12 longitudinal isolates, with the highest number of low-frequency mutations between 3-5% frequency. Seven candidate drug-resistance mutations across five evolutionary trajectories showed frequency shifts that correlated with changes in minimum inhibitory concentrations to the corresponding antibiotics. These included a 23S rRNA clarithromycin-resistance SNP detected at 7% frequency when phenotypic resistance emerged, suggesting that low-frequency variants drive subpopulation evolution. Acquisition of drug resistance during therapy was associated with several low-frequency mutations in genes associated with resistance to antibiotics, including clarithromycin and quinolones, in other NTM species.

**Conclusion:** This study highlights the importance of low-frequency variants as drivers of intra-patient bacterial population diversity, allowing subpopulations to adapt to antibiotic pressure and ultimately contributing to treatment failure. Additionally, it underscores their potential implications for the development of molecular diagnostic tests for NTM resistance prediction.

## Background

Nontuberculous mycobacteria (NTMs) are emerging pathogens that can infect both immunocompromised and immunocompetent hosts (1–6). These increasingly common pathogens pose several diagnostic and therapeutic challenges (7). Many species are resistant to multiple antimicrobial agents; treatment failure, emergent antimicrobial resistance and recurrent infection are frequent outcomes in clinical practice (8, 9). Approaches for genotypic identification and testing for antimicrobial drug susceptibility are not fully standardized, especially for less common species. Moreover, results from *in vitro* susceptibility assays correlate imperfectly (and sometimes not at all) with clinical outcomes (10–15). Failure to predict treatment effectiveness likely relates to several key factors: diversity of infecting bacterial populations within a host, suboptimal antimicrobial concentrations at privileged host sites, and unidentified bacterial adaptations that lead to persistence in the presence of antimicrobials (16–20). As a result, therapeutic failure is common in NTM treatment despite the use of antimicrobial agents that appear to be active *in vitro*.

Another significant challenge related to NTM diagnosis is the heterogeneity of methods used to determine species identity. Traditional high-performance liquid chromatography has been largely superseded by matrix-assisted laser desorption/ionization time-of-flight (MALDI-ToF) and genomic methods such as DNA probes and selective gene sequencing (e.g. the ribosomal 16S gene). However, different laboratories use different methods, and this has clinical implications for correct species assignment. Both MALDI-ToF and single-gene sequencing have been associated with species misclassification in clinical specimens (21), and sequencing of multiple genes or whole genome sequencing (WGS) are increasingly required for accurate NTM speciation.

In this study, we employed high-resolution genomics to characterize the intra-patient population dynamics of a disseminated infection caused by a previously uncharacterized NTM species, *Mycobacterium sp. SMC-2*. We combined long-read sequencing to construct a high-quality *de novo* assembly with longitudinal short-read WGS to track genomic variation across 12 serial isolates collected over the 29-month infection period. This dual-platform approach allowed us to examine the evolutionary trajectory of the bacteria during infection and in response to antibiotic therapy and identify the mechanisms driving phenotypic shifts under antibiotic pressure.

## Results

In order to understand the genetic basis of treatment failure in NTM disease, we analyzed bacterial genome dynamics across a prolonged course of antibiotic treatment. We examined longitudinally sampled isolates from a male in his 70s, who had presented >5 years after an orthotopic heart transplant for ischemic cardiomyopathy with approximately 4 months of fatigue, intermittent night sweats, and significant weight loss. Initial evaluation revealed hypercalcemia with associated acute renal failure as well as splenomegaly and upper abdominal/retroperitoneal lymphadenopathy on computed tomography. Lymph node and bone marrow biopsies demonstrated necrotizing granulomatous inflammation, and a blood culture grew an organism initially identified as *Mycobacterium nebraskense* by 16S rRNA target sequencing. A urine culture collected on day 18 of hospitalization grew the same organism, and on hospital day 19 therapy with azithromycin, rifabutin, and ethambutol was initiated.

After about 6 weeks of therapy he demonstrated clinical improvement, with decreased night sweats and weight gain. Several subsequent blood cultures continued to grow the same mycobacterium **(Figure 1)**. Initial pDST results **(Supplemental Table 1)** suggested *in vitro* susceptibility to numerous antimicrobials, and ethambutol was switched to doxycycline after about two months. The patient had continued clinical improvement despite ongoing positive blood cultures. 8.5 months after initial diagnosis, he remained clinically well but had a positive blood culture for which susceptibility testing demonstrated new resistance to clarithromycin. The regimen was switched to moxifloxacin, clofazimine, and doxycycline at month 13 (**Figure 1**). Blood and urine cultures over the next several months remained positive. At month 19 he had a negative blood culture but experienced progression of existing renal failure. Blood cultures obtained at months 21 and 22 were also negative and repeat computed tomography imaging demonstrated stable abdominal lymphadenopathy and splenomegaly with new endplate erosion of the L3/L4 vertebrae, concerning for osteomyelitis. The patient clinically declined over the next several months despite negative blood cultures at months 23-27. At month 29 after presentation, he was admitted to the hospital for progressive clinical deterioration. Blood culture at that time grew the same NTM species and pDST was repeated (**Table 1**). The patient was placed on hospice care and eventually died 31 months after the initial presentation.

**Figure 1:**
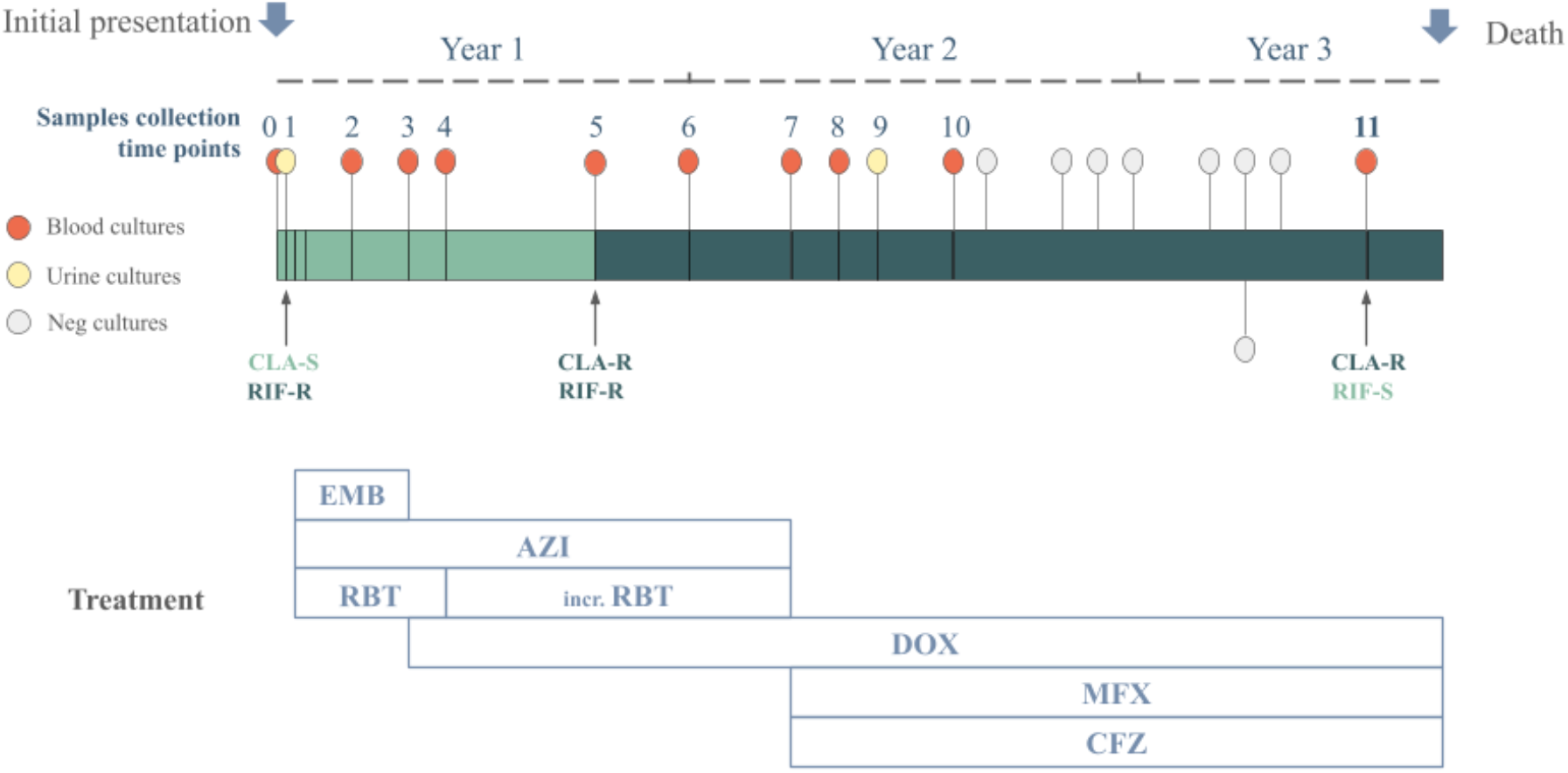
Summary of samples collection and treatment of the patient. Sample numbers correspond to all the samples sequenced. Yellow and red points correspond to positive urine and blood cultures, while white points correspond to negative cultures. Abbreviations: EMB-Ethambutol, AZI: azithromycin, RBT: rifabutin, inc RBT: increase of RBT dose, DOX, doxycycline, MFX: moxifloxacin, CFZ: clofazimine, RIF-R and RIF-S: rifampicin resistant and susceptible, CLA-R and CLA-S: clarithromycin resistance and susceptible.

**Table 1:**
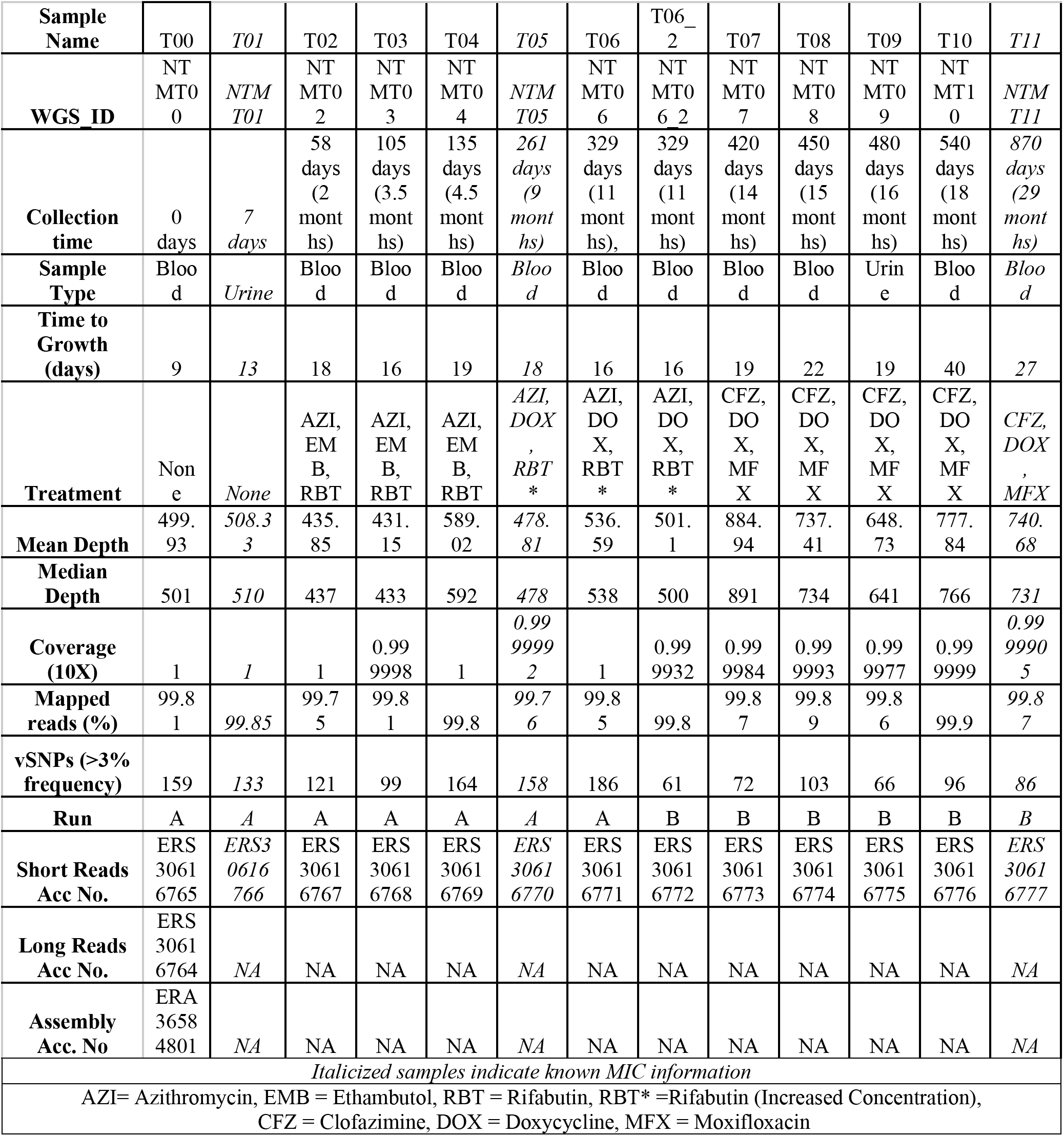
Sample Dictionary and Summary Statistics (Ordered by collection time)

Throughout the 29-month period, clinical isolates from several key timepoints had been archived. These included specimens from before the start of antibiotic treatment, samples from several timepoints where there was emerging antibiotic resistance, and samples during clinical recurrence. These strains and their corresponding genomes created a unique opportunity to examine infection trajectory, NTM population dynamics, and intra-host evolution.

### Identification of a rare NTM species through whole genome analysis

16S rRNA sequencing from a reference laboratory originally identified the strain as *M. nebraskense*, a NTM first described in clinical cases in 2006 (22) that is rarely associated with pulmonary disease in humans and has not been previously linked to disseminated infection (23). To obtain a baseline accurate genome for comparison, we sequenced the initial isolate (T00) by Nanopore long-read sequencing. The run obtained a 220,095,407 bp output, a total number of 75,400 reads, a sequencing depth of 37X and an N50 equal to 7,775 bp. Genome accuracy was estimated to be around Q60 (one error per 1,000,000 bases), allowing for genome assembly and bioinformatic analysis. The genome was resolved in 1 circular contig with 36X depth, a genome size of 5.9Mb, and 99.94% completeness. Genome annotation with Bakta predicted 5,528 genes including 5459 protein-coding (CDS), 39 non-coding RNA (ncRNA), 7 rRNA, 95 tRNA and 2 tmRNA. No plasmids were found.

Although the reference laboratory had originally identified the strain as *M. nebraskense*, the subsequent analysis of the assembled genome revealed a 99.95% average nucleotide identity (ANI) with the *Mycobacterium sp. SMC-2* genome (NCBI RefSeq accession GCF_025263485.1). In contrast, the ANI with *M. nebraskense* was below 95% (89.51%), suggesting that the NTM causing the infection was *M. sp. SMC-2* instead of *M. nebraskense.* This result was supported by a Mash distance-based tree, which confirmed *M. sp. SMC-2* as the closest genome available in the public databases **(Figure 2).**

**Figure 2:**
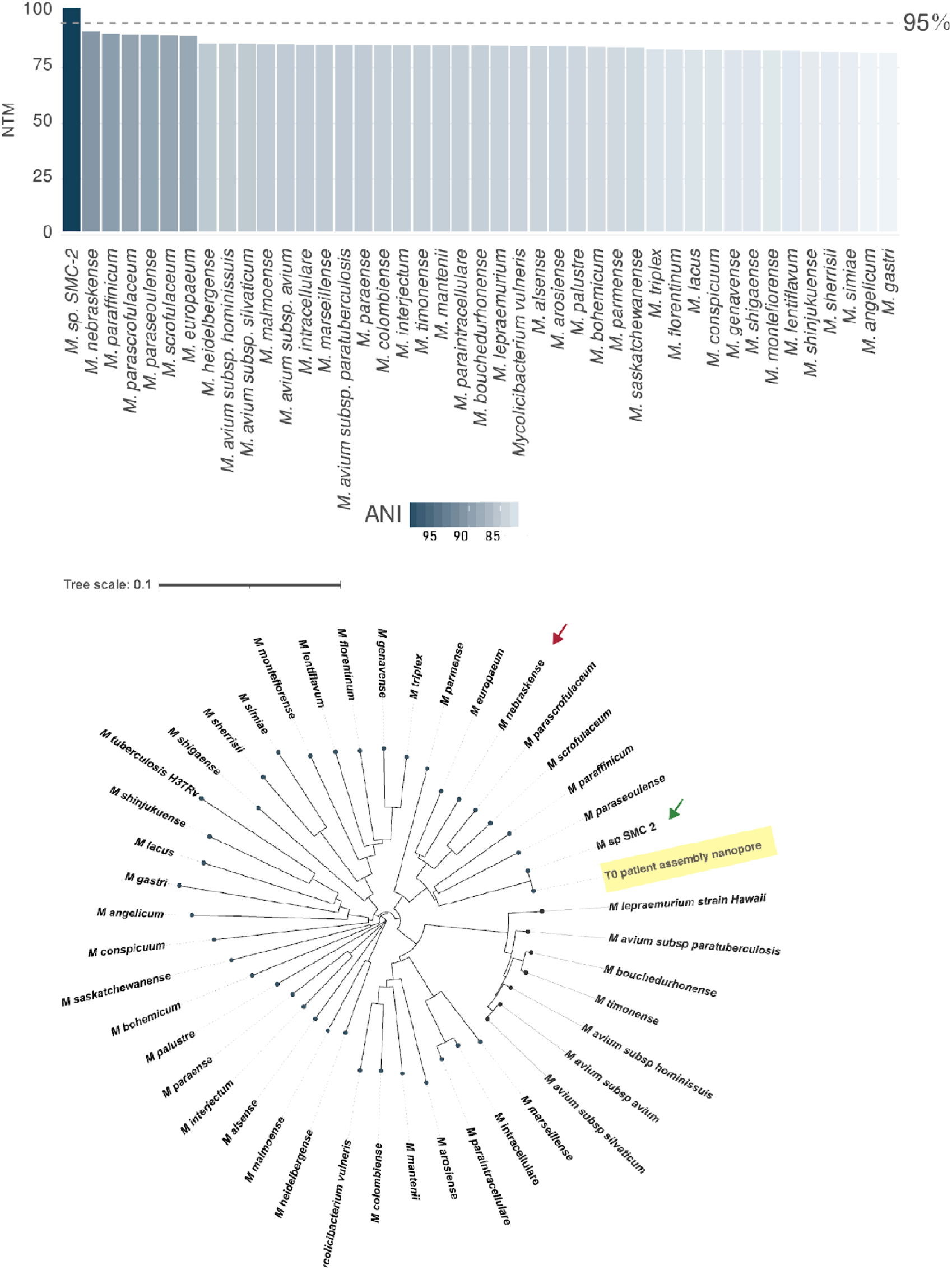
Genomic identification of the NTM. **(A)** ANI analysis comparing the T0_assembly with the 43 NTM genomes showing the highest genomic similarity. **(B)** Phylogenetic tree based on Mash distances for the same set of NTM genomes, with the NTM genome (T00) highlighted in yellow. Arrows highlight the *M. sp. SMC-2* (green) and *M. nebraskense* (red) genomes. Reduced plots display the 43 most closely related genomes out of a total of 175.

### Within host bacterial population dynamics

The T00 assembly was subsequently used as reference for mapping and variant analysis of the longitudinal short-read isolates. Samples were sequenced in two different runs achieving an average depth of 597.7X (range: 431.2X - 884.9X). In general, the mean depth was significantly higher in run B (t-test, p-value = 0.0073) (**Table 1, Supplemental Figure 1A**). All samples obtained more than 99.99% of the genome covered at 10X depth (**Table 1, Supplemental Figure 1A**).

Overall, the strict variant calling pipeline, designed to call variants as low as 3% frequency, enabled a robust analysis across all samples. Variant evaluation across all isolates identified 444 unique SNPs (130 synonymous, 248 non-synonymous, 3 in non-coding transcript regions, and 63 intergenic) and 26 indels (21 frameshift and 5 intergenic). The majority of these variants were detected at low frequencies (3%-10%). In addition, cultures sequenced in run A obtained a significantly higher number of ultra-low frequency variants (3%-5%) despite the lower depth compared to run B (mean run A: 44.7, mean run B: 23.3, p-value=0.0038) (**Supplemental Figure 1B**). Fixed variants (>90% frequency) were only observed in the final time points, T10 and T11, which carried 1 and 9 fixed SNPs, respectively **(Supplemental Figure 1B)**.

Comparison of variants among the isolates enabled the characterization of frequency dynamics across the longitudinal isolates. We filtered variants showing a frequency change of over 20%, obtaining a subset that comprised 36 SNPs and 8 indels. These mutations were classified into 10 groups according to their frequency shifts over time using the k-means algorithm, thereby suggesting a high population diversity (**Figure 3**).

**Figure 3:**
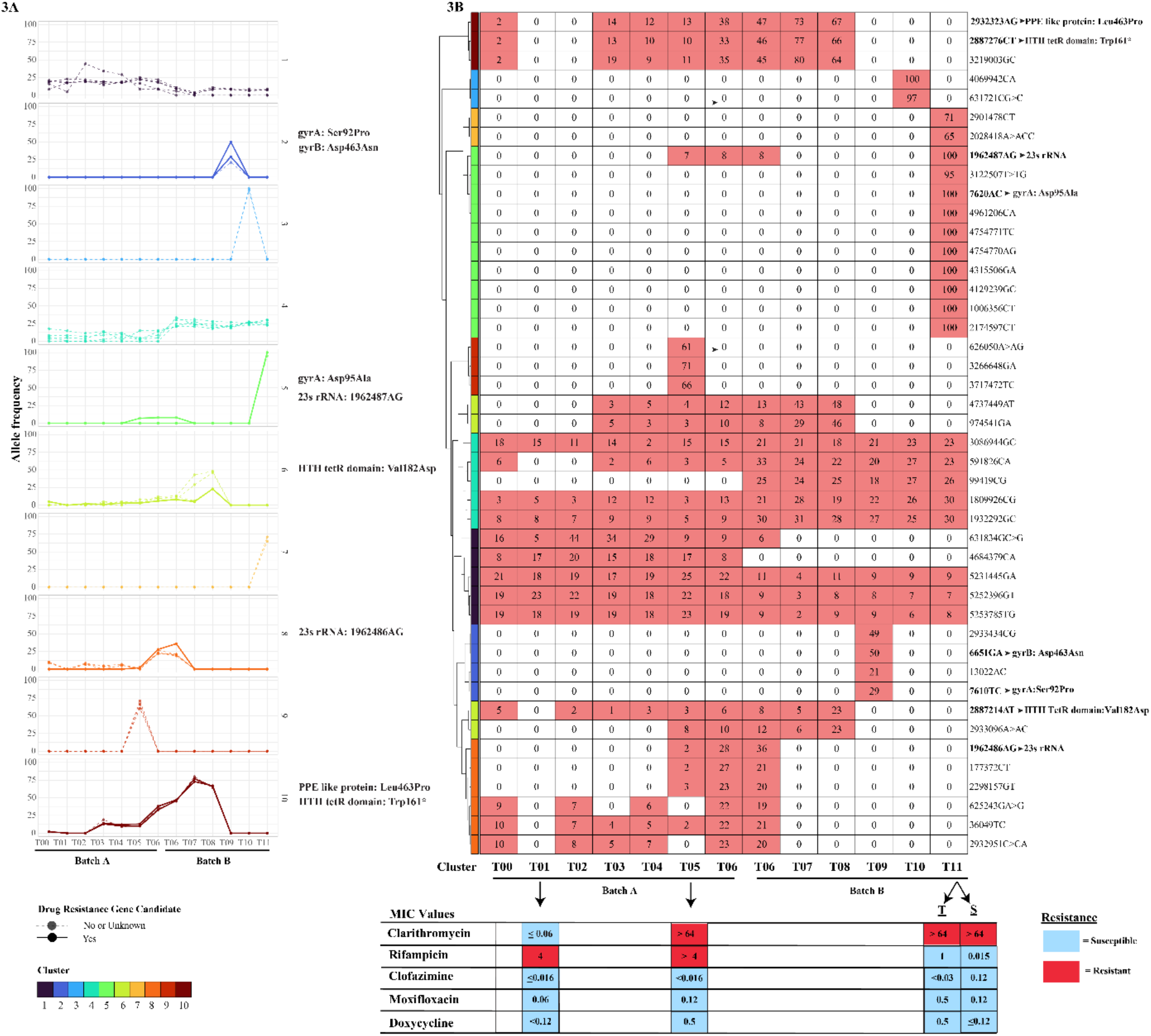
Analysis of SNP dynamics during treatment. **(A)** SNP frequency variation over time. Variants were clustered into 10 groups using the k-means algorithm based on similarities in their temporal frequency patterns. Solid lines represent candidate drug resistant associated mutations which are listed for each cluster on the right as well as bolded in **(B)** Presence/absense matrix of variant frequencies grouped by cluster. Numbers within each cell represent the variant frequency, and red highlights frequencies higher than 0. Drug-resistance candidates are highlighted in bold on the right side of the plot. Information on drug susceptibility and MICs to relevant antibiotics is provided below, with the light blue representing susceptibility to a drug and red representing resistance.

These dynamics were independently confirmed combining the Lolipop genotyping with Muller algorithms, which provided a high-resolution view of the subpopulation structure of the NTM infection. The Muller plot revealed high baseline population diversity, characterized by multiple co-existing clones at T00 which are represented by different colors in **Figure 4**. Most of the clones varied in frequency across samples, except those that were only present in individual timepoints. The Muller Plot indicated that most of the clones were present before treatment started, and their subsequent shifts during treatment suggested a complex clonal evolution pattern. Thus, we leveraged the diversity at baseline, defined as 12-SNPs distance to the last common ancestor [29], to estimate when the initial infection/colonization occurred. As there was no available molecular clock for this novel NTM species, we cautiously applied a MAC short-term substitution rate (2.05*10^-07^ SNPs/site/year [95%CI: 1.56*10^-7^-2.71*10^-7^] based on AM. Walsh *et. al* (24)). Using the formula explained in methods, the point estimate for the infection/colonization time was 10 years prior to diagnosis (time 0) but with a wide confidence interval (95%CI: 5.2-17.58 years).

**Figure 4:**
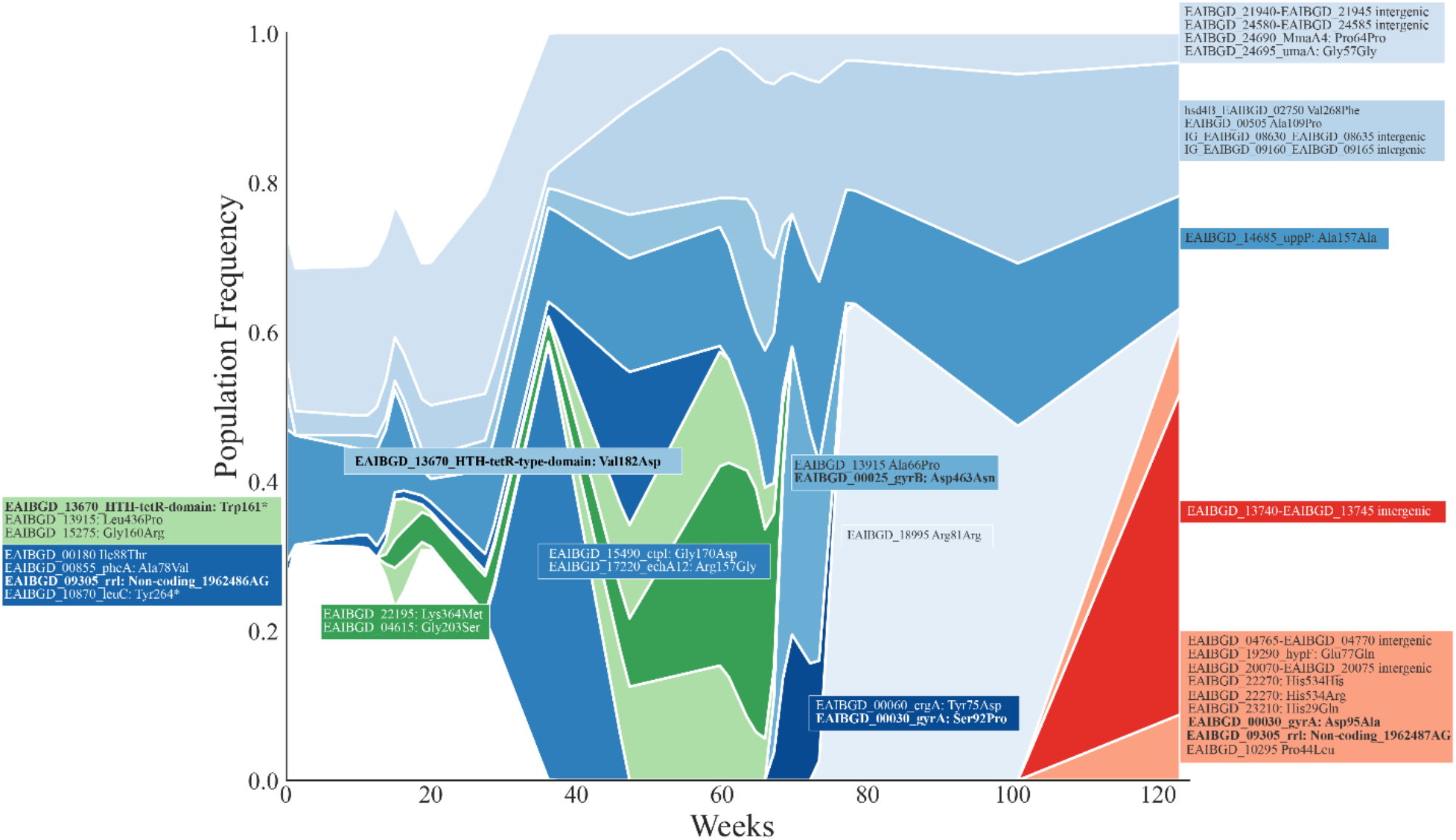
Muller plot representing the population dynamics during the 2-year infection period. Colors represent different genotypes. Labels inside the colored boxes represent new variants arising in each genotype, with bold labels highlighting candidate mutations in drug-resistance associated genes.

### Mutational trajectories identified antibiotic resistance driver mutations

The frequency trajectories of the variants, classified in 10 clusters by the k-means algorithm, suggested that either genetic drift, (e.g.: compartmentalization at different sites) or positive selection could act as the primary driver of these dynamics. As antimicrobials are a major selective pressure during treatment, we searched for putative drug-resistance conferring mutations potentially explaining these trajectories. To this end, we combined the annotation of SNPs assigned to each cluster with MIC values for relevant antimicrobials used during treatment. We focused on drug susceptibility tests showing changes in MIC over time. Based on these MIC shifts, we screened for variants in genes conferring resistance to rifampicin, fluoroquinolones, doxycycline and clarithromycin that could explain the observed changes. This analysis identified seven candidate SNPs: two located in the 23S rRNA, one in *gyrB*, two in *gyrA*, and two in a tetR-type domain–containing gene. Different candidate variants correlated with five distinct trajectories **(Figure 3A),** suggesting that they may act as putative drivers of subpopulation variation.

Clusters 5 and 8 included SNPs found in 23S rRNA (*rrl)* gene, responsible for macrolide (clarithromycin/azithromycin) resistance. 1962487AG (*rrl* position 2059, cluster 5) and 1962486AG (*rrl* position 2058, cluster 8) are located in the peptidyl transferase loop of the 23S rRNA gene, the V domain **(Supplemental Figure 2A**) and have both been previously identified as mutations associated with macrolide resistance (25–30). 1962487AG appeared at 7% frequency in T05, 8% in T06, and fixed at 100% in T11 **(Figure 3, Supplemental Figure 2D)**. These differences in SNP frequency were concordant with the MIC shift of clarithromycin **(Table 1)**. Similarly, 1962486AG was found in T05 at frequency 2%, peaked at T06 with frequency close to 30% and then disappeared **(Figure 3, Supplemental Figure 2D)**. These SNPs temporally correlated with the administration of azithromycin, which started after T01 and was discontinued at T06, with MIC data consistent with phenotypic resistance to clarithromycin at T05 and T11 (**Table 1**).

Clusters 2 and 5 also included SNPs likely associated with fluoroquinolone resistance. In cluster 2 there were two candidate mutations appearing only in T09. These two mutations were gyrA_Ser92Pro at 29% frequency and gyrB_Asp463Asn at 50% frequency. In cluster 5, gyrA_Asp95Gly was detected at 100% frequency at T11 **(Supplemental Figure 2D)**. Mutations found in *gyrA* were homologous to well-characterized drug resistance associated variants in other mycobacterial species and were located within the Quinolone Resistance-Determining Region (QRDR), in the N-terminal region of the GyrA subunit **(Supplemental Figure 2B)**. The comparative genomics analysis of GyrA amino acid sequence between this NTM and *Mycobacterium tuberculosis (Mtb)*, H37Rv strain, revealed that the corresponding mutations in *Mtb* are related to moxifloxacin and levofloxacin resistance. Specifically, these substitutions are listed in the WHO catalogue as high-confidence resistance associated variants in *Mtb* (31). The same analysis for *gyrB* candidates revealed that the mutation found in T09 (Asp463Asn at 50% frequency) is at the beginning of the QRDR region in GyrB, which has been documented in *in vitro* selected fluoroquinolone resistant strains, but limited association with resistance in *Mtb* clinical isolates (32–35) **(Supplemental Figures 2B and 2D)**. A fluoroquinolone (moxifloxacin) was added to treatment at month 13, between T06 and T07, and MIC values of the fluoroquinolone ciprofloxacin increased at T05 and T11 (**Supplemental Table S1**). Thus, the candidate mutations found explained the MIC increase at T11. However, we have not found a mutation explaining the increased MIC values at T05.

With regard to doxycycline resistance, further analysis failed to identify specific candidate mutations. HTH TetR domains were predicted in several coding regions, and mutations in those domains were found in clusters 6 and 10. Cluster 6 mutations peaked at T08 and were transient, while cluster 10 mutations involved a stop codon and were present at T03, peaking at T07, and then disappearing. The temporal change in these mutation clusters did not correlate with doxycycline administration starting at T03 and subsequent MIC data showing an increase in MIC at T05, which was maintained at T11.

To further assess potential mutations driving antibiotic resistance, we focused on Cluster 10, where the pattern of mutations followed the MIC trends for both rifampicin and clarithromycin. Briefly, the largest drivers of Cluster 10 were SNP 2887276CT (Trp161*) at the HTH tetR domain above described and SNP 2932323AG (Leu436Pro) at a PPE like protein. These SNPs appeared at low frequency in T00 and T05 (2% and 10% respectively), when the strain was considered resistant to rifampicin by phenotypic drug susceptibility testing (pDST). The frequency of these SNPs increased during the period of rifabutin (a rifamycin) usage, reaching almost 60% by the time rifabutin treatment was stopped at T06, and subsequently dropped back down to 0% when the strain was rifampicin sensitive according to MIC data **(Supplemental Figure 3)**.

Despite the MIC phenotypes in T01 and T05 being classified as rifampicin resistant, no variants were found in the *rpoB* gene. In fact, the rifampicin MIC was higher in the T00 isolate compared with the T11 isolate, suggesting that rifampicin resistance mutations might have been present at the initial timepoint. To further analyze this, the RpoB amino acid sequence from the NTM baseline T00 genome assembly was aligned to that of *Mtb* H37Rv. This approach aimed to identify baseline homologous SNPs and indels corresponding to resistance-associated variants reported in the WHO catalogue (2023 version) (36). The results showed 52 non-synonymous SNPs, one 5-amino acid insertion, and two deletions (25 and 3 amino acids) in the T00 assembly **(Supplemental Table S2).** However, none of the identified mutations were listed in the WHO catalogue of drug resistance in *Mtb* (36). Additionally, we confirmed the absence of the *arr* gene, which is responsible for rifampicin inactivation in other mycobacterial species such as *M. abscessus*.

The drivers for the remaining mutational trajectories were not clear as we failed to identify likely candidate mutations. However, clusters 3, 7 and 9 involved only one time point, while cluster 4 involved a series of mutations present in all samples albeit increasing frequency after a major change in treatment at T06, thus unlikely to involve antimicrobial resistance.

## Discussion

This in-depth analysis of temporal genomic variation of an atypical NTM during disseminated infection in an immunocompromised patient revealed several unexpected insights. First, the initial species identification, obtained by single-gene sequencing, was incorrect. While the patient was initially diagnosed with a highly atypical, disseminated *M. nebraskense* infection, the causative agent was actually an even rarer organism provisionally named *M. sp. SMC-2*. Second, despite the presence of ongoing antimicrobial therapy and development of phenotypic resistance, drug-resistance mutations were only present at low frequencies. Third, assuming that the initial infection was monoclonal and using reasonable mutation rate estimates suggested either that infection might have reasonably occurred prior to immunosuppression or that the initial infection occurred after immunosuppression but was polyclonal. All three of these insights bear further discussion.

This study corroborated prior work on other NTM demonstrating that using only one gene for species identification may lead to errors (21). Other commonly used identification techniques such as MALDI-ToF may also misclassify NTM species (37). In this case, a strain that was diagnosed as *M. nebraskense* from 16S rRNA sequencing turned out to be a different NTM species. What was initially thought to be a heretofore undescribed manifestation of a known organism, turned out to be a disseminated infection with a different, even rarer organism. This species, identified as *M. sp* SMC-2, has only a single genome available in public databases, complicating efforts to understand the significance of mutations that emerged during therapy. Many phenotypic and genomic analyses that are relatively “standard” for example in *Mtb* (e.g.: MICs, genetic diversity, and heteroresistance), have not been optimized for the majority of NTM (10–15). As WGS becomes increasingly used as part of clinical microbiology practice, more atypical or heretofore unidentified NTM species are likely to be identified, and this will present challenges related to choice of empiric therapy and interpretation of antimicrobial susceptibility results for these rare species.

One of the major challenges when dealing with unusual NTM infections such as the one described here is the lack of an appropriate reference genome. In the case of *M. sp. SMC-2*, only a single published reference genome was available for comparison, and it was not fully annotated. This required *de novo* annotation of the genome, both to identify the presence or absence of antibiotic resistance genes and to identify likely candidate resistance mutations. Annotation of candidate resistance mutations for clarithromycin and quinolones was straightforward, as they occur in highly conserved genes with homologous positions in other bacteria. In contrast, for doxycycline we can only postulate candidate mutations in predicted HTH TetR domains, which are difficult to interpret without additional experimental validation.

Given prolonged azithromycin-based treatment and phenotypic change from susceptibility with a low clarithromycin MIC to highly resistant with a high clarithromycin MIC, we would have expected to find acquisition of a high-frequency mutation in the 23S ribosomal RNA gene, similar to what is typically found when MAC complex organisms develop macrolide resistance (38). However, we only found this mutation at low frequency (7%) at the timepoint at which clarithromycin resistance was initially detected (and at which time the patient was receiving azithromycin). The low frequency of this mutation when the patient was actively receiving drug is unexplained, although perhaps could relate to the relatively low levels of azithromycin reported in plasma and some other tissue compartments (the drug concentrates intracellularly, particularly in white blood cells) (39). While heteroresistance is well-described in *Mtb*, it is less understood in NTM. However, at least one study (40) clearly demonstrated heteroresistance to macrolides (similar to our findings) in MAC that could often only be detected when the organism was subcultured in a clarithromycin-containing medium. High-depth genomic sequencing of the clinical NTM strain revealed the importance of these mixed populations in conferring antibiotic resistance. In fact, we were able to identify a driver mutation linked to antibiotic resistance in at least five out of the ten trajectories. Our data thus suggest that antibiotic pressures were major drivers of diversity within the patient over time. We found that low frequency SNPs (<10%) appeared in genes with known contribution to resistance, including 23S rRNA, *gyrA*, and *gyrB*. The appearance of these mutations correlated with their respective antibiotic usage as well as MIC readings. In *Mtb*, there is an ongoing discussion about the level at which heteroresistance is clinically relevant, particularly given combined treatments (41–46), and when a change in antibiotics and/or regimen is advisable. Our data support the idea that very low-frequency variants exist and drive changes in MIC, and although these data come from a single patient, our findings indicate an impact on clinical evolution, likely reflecting poorer regimen efficacy in NTM infections compared with more standardized tuberculosis treatments. These findings are also relevant to development of molecular assays to detect drug resistance in NTM; if low-frequency heteroresistance is driving phenotypic resistance, molecular assays will need to be able to detect these low-frequency variants to have adequate sensitivity for clinical use.

Our findings with regard to the association between other low-frequency mutations and phenotypic drug resistance are less clear. However, we did note some correlation between phenotypic MICs and the presence of low-frequency mutations in genes likely to be associated with drug resistance. For example, Cluster 2 showed mutations that arose after a drug regimen change to moxifloxacin, clofazimine, and doxycycline. Specifically, we identified a mutation in *gyrA* (Ser92Pro) and one in *gyrB* (Asp463Asn), which are both known to confer resistance to fluoroquinolones such as ciprofloxacin and moxifloxacin (33, 34). These mutations were detected at low frequency in T09, two months after the patient was exposed to moxifloxacin, but were absent in all other isolates. Additionally, a different *gyrA* mutation, Asp95Ala, was found exclusively in the final sample, T11, where it was fixed. This suggests the presence of different subpopulations under selective pressure from antibiotics. Although the MIC values for moxifloxacin and ciprofloxacin did not reach the formal clinical breakpoints established for *M. kansasii* (47), a notable MIC shift was observed following the emergence of *gyrA* and *gyrB* mutations. This reduced susceptibility, correlating with mutations in the QRDR, reiterates the limitations of current criteria to interpret clinical breakpoints when the species is unknown. Our approach should be viewed as hypothesis-generating: The described mutations are well known in homologous genes in other mycobacteria and are associated with resistance (particularly those in *gyrA*) (48–52). Further experiments including phenotypic testing of NTM enriched for these particular mutations and *in vivo* testing of the relevance of the mutations in different antibiotic regimens are warranted.

We combined long- and short-read sequencing to understand the diversity of *M. sp.* SMC-2 at the onset and during treatment. Our approach enabled recording the emergence of variants as low as 3% in the culture population, thus recording variation at higher resolution than the standard genomic analysis. The diversity observed at baseline (T00) is striking, as we identified many mutations that remained months, or even years, after treatment initiation. This suggests either that the infection had been established for some time or that the initial inoculum consisted of a large, diverse population. The latter is plausible, as many environmental mycobacteria harbor significant genetic diversity, and environmental exposures are likely to result in concurrent infections with multiple related organisms. Our preliminary analysis, using molecular clock estimates from MAC as a proxy, suggests that if the initial infection were monoclonal, the patient’s initial infection would have occurred between 5.2 and 17.6 years before initial presentation, while immunosuppressive therapy had only been initiated 6 years prior to initial presentation. While limited published data suggests latency of other NTM infections in animal models, (53, 54), a more likely scenario is that polyclonal infection occurred in our patient after the heart transplant. Even though a precise date cannot be determined in the absence of a species-specific molecular clock, our findings demonstrate the potential of genomic analysis to reconstruct the timeline of infection/colonization. This type of analysis could potentially be useful for other NTM infections, as understanding the time when infection occurred has significant implications in considering interventions to mitigate environmental exposure to NTM.

Our analysis suffers from several limitations. First, we used frozen subcultures of the NTM organism for analysis, which may not have fully represented the diversity of the population of organisms infecting the patient at each timepoint. Second, there are no clinical standards for phenotypic testing and clinically relevant antibiotic concentration cutoffs of this particular NTM; interpretations of susceptibility/resistance were extrapolated from other NTM. Third, the mutation rate in this particular NTM is unknown; using the mutation rate of another slow-growing NTM (MAC) as an estimate seemed reasonable but may have biased the calculations of time from the time of initial infection.

## Conclusions

This analysis demonstrates the complexities surrounding NTM identification and antimicrobial susceptibility testing. It reinforces the message that whole-genome sequencing, or at least sequencing of multiple genes, is essential for accurate NTM speciation. Furthermore, this study demonstrates the importance of very-low frequency mutations that seem to be associated with drug resistance and how they can be informative for potential development of molecular diagnostic tests for NTM resistance prediction. Indeed, tests that are not very sensitive to low-frequency populations will be likely to miss these clinically important mutations, as would sequencing with relatively low-depth of coverage. While selection for groups of mutations associated with drug therapies suggests that those drugs have antimicrobial activity, that effect is incomplete and poorly reflected by standard *in vitro* testing. Therefore, further longitudinal studies that employ deep-sequencing of sequential, within-patient isolates over time (such as in pulmonary MAC) may help to clarify the connection between standard *in vitro* phenotypic susceptibility testing and molecular markers of resistance.

## Methods

### Sampling Methods

Blood and urine specimens were obtained from the patient for clinical purposes at different timepoints as noted above. When NTM growth occurred, colonies were scraped from a subculture grown on Lowenstein-Jensen media and frozen at −80°C. Frozen subcultures were thawed and inoculated in 7H9 liquid media at 37°C. DNA was subsequently extracted and sent for short-read deep-sequencing using the NextSeq 1000 Illumina platform with the P1 XLEAP reagent kit (300 cycles). In total, 12 samples (T00-T11) were sequenced in 2 different runs, with sample T06 sequenced in both runs as an internal control after reculturing (**Table 1**).

### Phenotypic drug susceptibility testing

Minimum inhibitory concentration (MIC) to different drugs was assessed at different time points: T01 (week 1), T05 (week 36) and T11 (week 124). Drugs tested included trimethoprim-sulfamethoxazole, doxycycline, linezolid, rifabutin, amikacin, moxifloxacin, clofazimine, ciprofloxacin, clarithromycin, minocycline, rifampicin, bedaquiline, omadacycline, eravacycline and tedizolid. Susceptibility testing was done at a reference laboratory (The Mycobacteria/Nocardia Laboratory at the University of Texas-Tyler) via microdilution using the standard CLSI method (47). Cutoffs for phenotypic resistance prediction were determined using the standard for drug resistant *M. kansasii*. Due to the lack of standardized cutoffs, resistance could not be assessed for clofazimine, omadacycline, bedaquiline, eravacycline, and tedizolid; therefore only MICs were reported for these drugs (**Supplemental Table S1**).

### Identification methods

The NTM species causing the infection was identified by using two different methodologies. A PCR amplification of a 552 base pair segment of the 16S ribosomal RNA gene followed by sequencing was performed in the first two isolates through Mayo Clinic (55) and UT Tyler Galveston (56) labs, and WGS of the first isolate (T00). WGS was performed using both Oxford Nanopore PromethION long-read and Illumina short-read platforms through the company Plasmidsaurus. Briefly the library for T00 long-read sequencing was constructed using the v14 library prep chemistry from Oxford Nanopore and sequenced in a R10.4.1 flowcell. Basecalling was done using Dorado v4.3 Supper-Accurate model with Q10 filtering. Flitlong v0.2.1 was used for the quality filtering step, removing the worst 5% reads. The genome assembly was generated with Autocycler with three different assemblers (Flye v.2.9.6+, Hifiasm, and Plassembler v1.8.0+), and it was subsequently polished with Illumina short-reads to improve the quality using Medaka (v1.8.0). Annotation of the genome was done via Bakta v1.11. Assembly quality was evaluated by analyzing contigs with Bandage v0.8.1 and assessing completeness and contamination with CheckM v1.2.2. (57, 58). Finally, the genome was recircularized with circlator v1.5.5, fixing the start of the genome at the *dnaA* gene (59). The T00 assembly starting with *dnaA* gene was re-annotated with Backta, and Eggnogg (DIAMOND algorithm) was used to find gene orthologs in other bacterial species (60). Strain identification was performed through a whole genome comparison of the T00 assembly with 175 NTM genomes using an average nucleotide identity (ANI) program (61, 62). Phylogeny was then determined based on MASH distances [20]. Additionally, plasmid screening was performed using the PHASTEST online tool (63). The T00 assembly was also used as a reference genome for subsequent analyses. (64).

### Bioinformatic Analysis Methods

Illumina short-reads were filtered with fastp v0.23.2 (65) and mapped with BWA-mem (v0.7.17) (65) against the nanopore assembly obtained from the first isolate (T00). Genome coverage and depth was obtained using the tool bamqc included in QualiMap (v2.2.1)(66). To obtain high confidence single nucleotide polymorphisms (SNPs), we applied three different variant callers (Mutect2 (67), LoFreq (68) and VarScan2 (69)) and generated a consensus of all the variants called at least by 2 callers. We also performed a filtering step to keep the variants supported by two strands, with >3% allele frequency and >20X sequencing depth. Insertions and deletions (indels) were called only by LoFreq, to ensure high indel accuracy, and filtered considering mapping quality above 60, minimum depth 30X, frequency 3% and strand bias.

Intrapatient diversity was analyzed by comparing all the variants across all the isolates by using a customized Python script (70). A rescue step was included to recover the variants that did not pass depth and frequency filtering cut-offs. The frequency trajectories of the variants across the isolates were clustered using the k-means algorithm (with the Gap statistic method)(71, 72). While all variants were used to assess genetic diversity within a sample, for drug-resistance candidates, trajectories were considered relevant if the difference between the maximum and minimum frequency between the isolates exceeded 20%. Additionally, mycobacterial population dynamics were also assessed with the Lolipop package (v0.9.0) (73) by generating a Muller plot to visualize the abundance and genealogy of the estimated genotypes throughout the course of the infection.

We identified variants in drug-resistance associated genes for the antibiotics showing MIC shifts (*gyrA, gyrB, rpoB* and 23S rRNA or *rrl*). To determine if candidate mutations had been previously described as resistance-associated in other species, a comprehensive comparative analysis was performed. Specifically, genes extracted from the T00 assembly were aligned against their orthologs in *Escherichia coli* (*E. coli*) or *M. tuberculosis* (*Mtb*) H37Rv strain to obtain the gene coordinate correlation.

In addition, the theoretical time to colonization was estimated by calculating the distance between the T00 assembly and the last common ancestor (LCA), assuming that the sum of variant frequencies within a population is equivalent to the average number of mutations per cell, as described by T. Lieberman *et al* (74) In the absence of species-specific estimates for the NTM under study, the estimated molecular clock of the *M. avium* complex (MAC) was used (24). Time to colonization (T) was derived from the formula d = μ*T*L where, d represents the LCA distance, μ is the substitution rate (SNPs/site/year), and L the length of the NTM genome (75).

The T00 assembly (ERA36584801) was used as a reference genome for subsequent analyses. All raw sequencing data (assembly, long reads, and short reads) have been uploaded to the ENA repository under the project accession number PRJEB115262. Samples’ accession numbers are listed in **Table 1**. The datasets as well as the scripts used for analyses and graphing are available on Zenodo (Project name: NTM dataset; DOI: https://www.doi.org/10.5281/zenodo.21339451).

## Data Availability

All data produced are available online at Zenodo (https://www.doi.org/10.5281/zenodo.21339451)

## Declarations

### Ethics approval and consent to participate

Use of the patient clinical data and stored mycobacterial isolates was approved by the Duke Institutional Review Board (Protocol Pro00107795 and Pro00113019) including signed informed consent from the patient.

### Consent for publication

Not applicable.

### Availability of data and materials

The datasets generated and analyzed during the current study are available in the ENA repository, under the project accession number PRJEB115262. The datasets generated as well as the scripts used for analyses and graphing are available on Zenodo (https://www.doi.org/10.5281/zenodo.21339451).

### Authors’ contributions

J.E.S., I.C., A.R.M., C.M.L. and D.M.T. conceived and designed the project. E.K.M, N.A. and M.J.L. collected the samples and provided relevant case and strain information. A.X.M. cultured samples and extracted DNA from each timepoint. C.M.L, A.R.M, and M.G.L carried out the bioinformatic data analysis. I.C, J.E.S, D.M.T., A.R.M, M.G.L and C.M.L interpreted data and drafted the manuscript with input from all authors. All authors reviewed, edited, and approved the final manuscript.

## Acknowledgements

We thank Dr. Qingyun Liu and Dr. Mingyu Gan for assistance with bioinformatic identification of the initial strain.

## SUPPLEMENTAL INFORMATION

**Supplemental Table S1:** MIC of the isolates collected at different timepoints during treatment. MIC was performed by microdilution per CLSI standards (*M. kansasii*) at a reference laboratory.

| Isolate | T01 | T05 | T11_Transparent | T11_Smooth |
| --- | --- | --- | --- | --- |
| Week | 1 | 36 | 124 | 124 |
| Site | Urine | Blood | Blood | Blood |
| Antibiotic regimen | None | AZI, DOX, RBT* | CFZ, DOX, MFX | CFZ, DOX, MFX |
| Amikacin | S (MIC ≤1) | S (MIC ≤1) | S (MIC ≤1) | S (MIC ≤1) |
| Bedaquiline | N/A | U (MIC 0.004) | U (MIC 0.03) | U (MIC 0.008) |
| Clarithromycin | S (MIC ≤0.06) | R (MIC >64) | R (MIC >64) | R (MIC >64) |
| Clofazimine | S (MIC ≤0.016 ) | S (MIC <0.016 ) | S (MIC 0.03 ) | S (MIC 0.12) |
| Ciprofloxacin | S (MIC ≤0.12) | S (MIC ≤0.12) | S (MIC 2) | S (MIC 1) |
| Doxycycline | S (MIC <0.12) | S (MIC 0.5) | S (MIC 0.5) | S (MIC ≤0.12) |
| Eravacycline | N/A | U (MIC 8) | U (MIC 0.12) | U (MIC ≤0.015) |
| Linezolid | S (MIC ≤1) | S (MIC ≤1) | S (MIC ≤1) | S (MIC ≤1) |
| Minocycline | I (MIC 2) | I (MIC 2) | I (MIC 1) | S (MIC 0.12) |
| Moxifloxacin | S (MIC 0.06) | S (MIC 0.12) | S (MIC 0.5) | S (MIC 0.12) |
| Omadacycline | N/A | U (MIC 8) | U (MIC 0.5) | U (MIC ≤0.015) |
| Trimethoprim-sulfamethoxazole | S (MIC ≤0.25/4.75) | S (MIC ≤0.25/4.75) | S (MIC ≤0.25/4.75) | S (MIC ≤0.25/4.75) |
| Rifabutin | S (MIC ≤0.12) | S (MIC ≤0.12) | S (MIC ≤0.12) | S (MIC ≤0.12) |
| Rifampicin | R (MIC 4) | R (MIC >4) | S (MIC 1) | S (MIC 0.015) |
| Tedizolid | N/A | U (MIC ≤0.25) | U (MIC 0.5) | U (MIC ≤0.25) |
| Resistance: S= Susceptible, R = Resistant, I = Intermediate, U= unknown |  |  |  |  |
| AZI= Azithromycin, EMB = Ethambutol, RBT = Rifabutin, RBT* =Rifabutin (Increased Concentration), CFZ = Clofazimine, DOX = Doxycycline, MFX = Moxifloxacin |  |  |  |  |

**Supplemental Table S2:** List of *rpoB* mutations.

| Alignment<br>POS | REF H37RV<br>POS | REF HR37RV<br>AA | QRY NTM_T00<br>POS | QRY NTM_T00<br>AA | SNP<br>INDEL |
| --- | --- | --- | --- | --- | --- |
| 1 | 1 | L | - | - | DEL |
| 2 | 2 | A | - | - | DEL |
| 3 | 3 | D | - | - | DEL |
| 4 | 4 | S | - | - | DEL |
| 5 | 5 | R | - | - | DEL |
| 6 | 6 | Q | - | - | DEL |
| 7 | 7 | S | - | - | DEL |
| 8 | 8 | K | - | - | DEL |
| 9 | 9 | T | - | - | DEL |
| 10 | 10 | A | - | - | DEL |
| 11 | 11 | A | - | - | DEL |
| 12 | 12 | S | - | - | DEL |
| 13 | 13 | P | - | - | DEL |
| 14 | 14 | S | - | - | DEL |
| 15 | 15 | P | - | - | DEL |
| 16 | 16 | S | - | - | DEL |
| 17 | 17 | R | - | - | DEL |
| 18 | 18 | P | - | - | DEL |
| 19 | 19 | Q | - | - | DEL |
| 20 | 20 | S | - | - | DEL |
| 21 | 21 | S | - | - | DEL |
| 22 | 22 | S | - | - | DEL |
| 23 | 23 | N | - | - | DEL |
| 24 | 24 | N | - | - | DEL |
| 25 | 25 | S | - | - | DEL |
| 52 | 52 | T | 27 | I | SNP |
| 67 | 67 | S | 42 | A | SNP |
| 70 | 70 | E | 45 | G | SNP |
| 73 | 73 | D | 48 | E | SNP |
| 74 | 74 | V | 49 | P | SNP |
| 75 | 75 | N | 50 | T | SNP |
| 108 | 108 | D | 83 | E | SNP |
| 190 | 190 | D | 165 | E | SNP |
| 192 | 192 | T | 167 | L | SNP |
| 239 | 239 | S | 214 | N | SNP |
| 243 | 243 | V | 218 | H | SNP |
| 253 | 253 | R | 228 | M | SNP |
| 254 | 254 | S | 229 | G | SNP |
| 262 | 262 | V | 237 | A | SNP |
| 318 | 318 | V | 293 | A | SNP |
| 344 | 344 | E | 319 | H | SNP |
| 345 | 345 | G | 320 | A | SNP |
| 346 | 346 | Q | 321 | R | SNP |
| 353 | 348 | T | 328 | V | SNP |
| 524 | 519 | S | 499 | T | SNP |
| 528 | 523 | V | 503 | H | SNP |
| 549 | 544 | A | 524 | G | SNP |
| 550 | 545 | D | 525 | K | SNP |
| 554 | 549 | V | 529 | E | SNP |
| 556 | 551 | P | 531 | S | SNP |
| 644 | 639 | E | 619 | D | SNP |
| 645 | 640 | E | 620 | K | SNP |
| 646 | 641 | S | 621 | A | SNP |
| 661 | 656 | H | 636 | A | SNP |
| 663 | 658 | N | 638 | D | SNP |
| 667 | 662 | R | 642 | H | SNP |
| 675 | 670 | A | 650 | E | SNP |
| 686 | 681 | C | 661 | S | SNP |
| 708 | 703 | D | 683 | E | SNP |
| 709 | 704 | D | 684 | N | SNP |
| 775 | 770 | I | 750 | V | SNP |
| 857 | 852 | E | 832 | D | SNP |
| 900 | 895 | V | 875 | Q | SNP |
| 907 | 902 | A | 882 | P | SNP |
| 947 | 942 | A | - | - | DEL |
| 948 | 943 | A | - | - | DEL |
| 949 | 944 | K | - | - | DEL |
| 951 | 946 | V | 923 | K | SNP |
| 953 | 948 | D | 925 | E | SNP |
| 957 | 952 | R | 929 | N | SNP |
| 960 | 955 | D | 932 | K | SNP |
| 964 | 959 | E | 936 | Q | SNP |
| 966 | 961 | Q | 938 | E | SNP |
| 980 | 975 | Q | 952 | R | SNP |
| 990 | 985 | C | 962 | A | SNP |
| 997 | 992 | G | 969 | N | SNP |
| 998 | 993 | D | 970 | E | SNP |
| 1000 | 995 | L | 972 | M | SNP |
| 1003 | 998 | A | 975 | G | SNP |
| 1008 | 1003 | M | 980 | V | SNP |

**Supplemental Figure 1:**
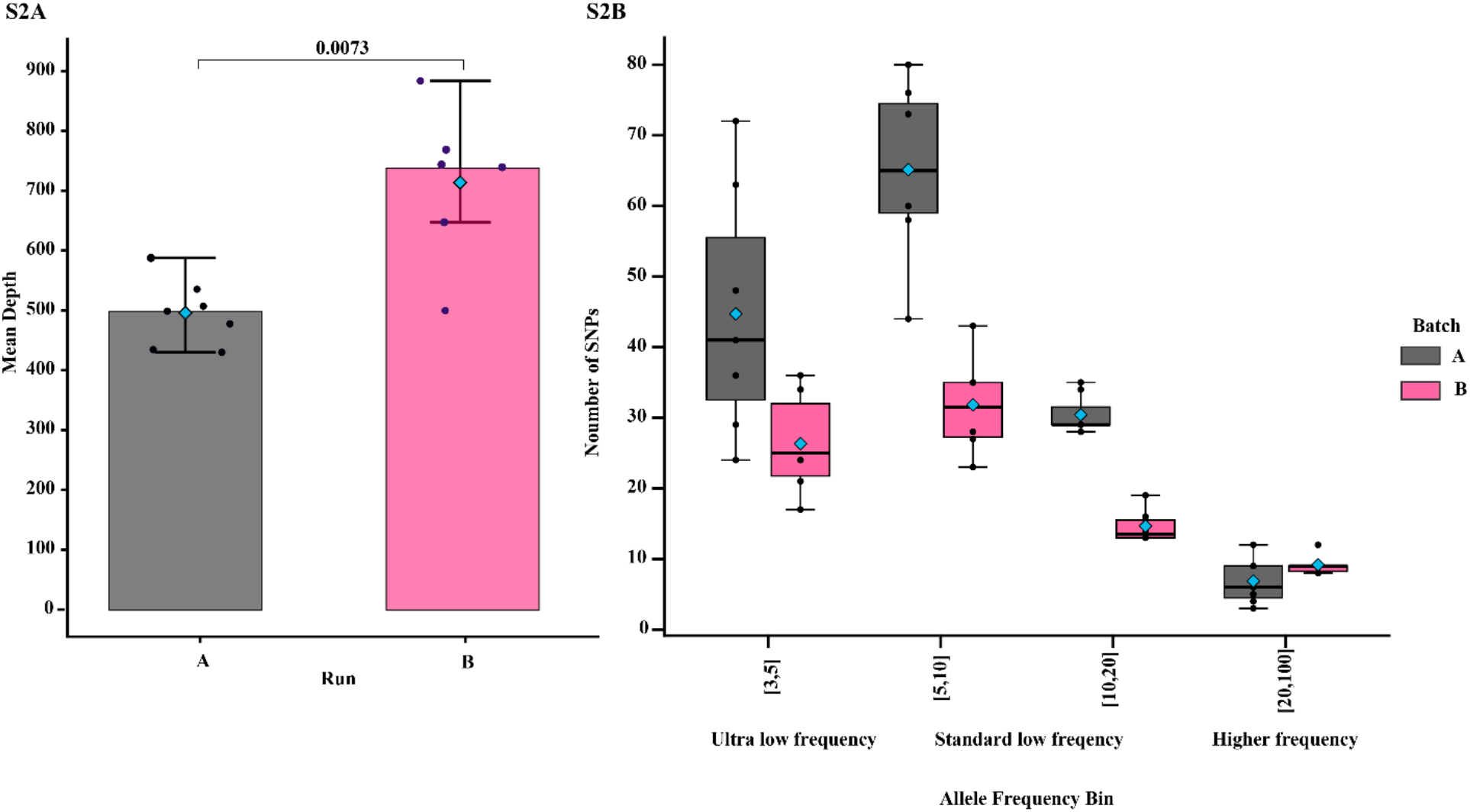
Sequencing results. **(A)** Comparison of mean depth obtained in each sequencing run while **(B)** compares the distribution of SNPs that appear at different frequencies between the two runs. Colors indicate the sequencing run, with light blue diamonds showing the mean.

**Supplemental Figure 2:**
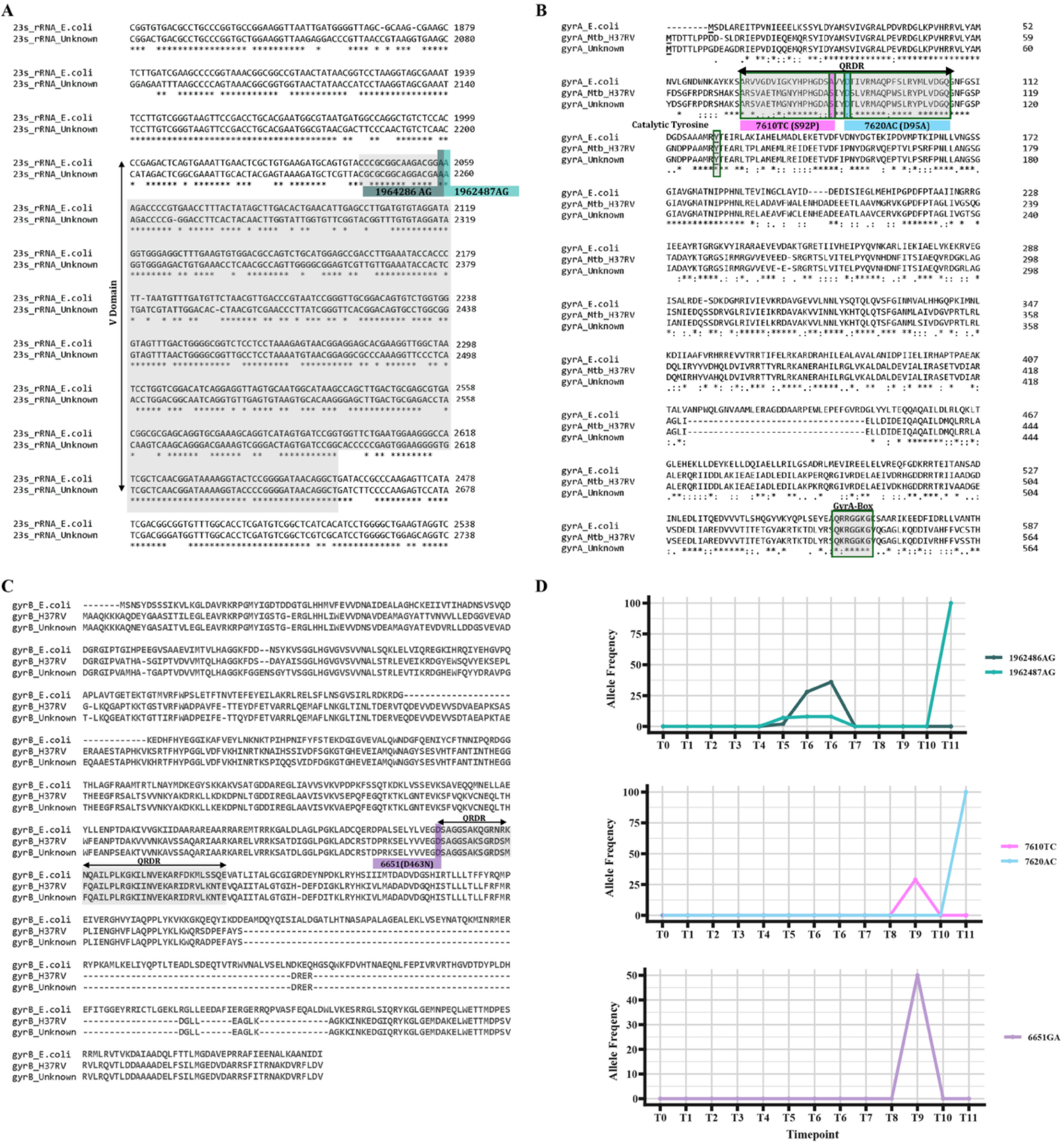
Analysis of drug-resistance conferring SNPs. **(A)** Alignment of V domain of the 23S rRNA nucleotide sequences of *E. coli* strain K12 and the T0 assembly, highlighting the positional correspondence of the mutations identified in the T00 isolate assembly. **(B and C)** Alignment of the *E. coli* and *Mtb* GyrA and GryB amino acid sequence to the T00 assembly highlight the SNPs that fall in the quinolone resistance determining region (QRDR). Dynamics of all candidate SNPs related to resistance are shown in **(D)**, and colors correspond to the SNPs highlighted in A-C.

**Supplemental Figure 3:**
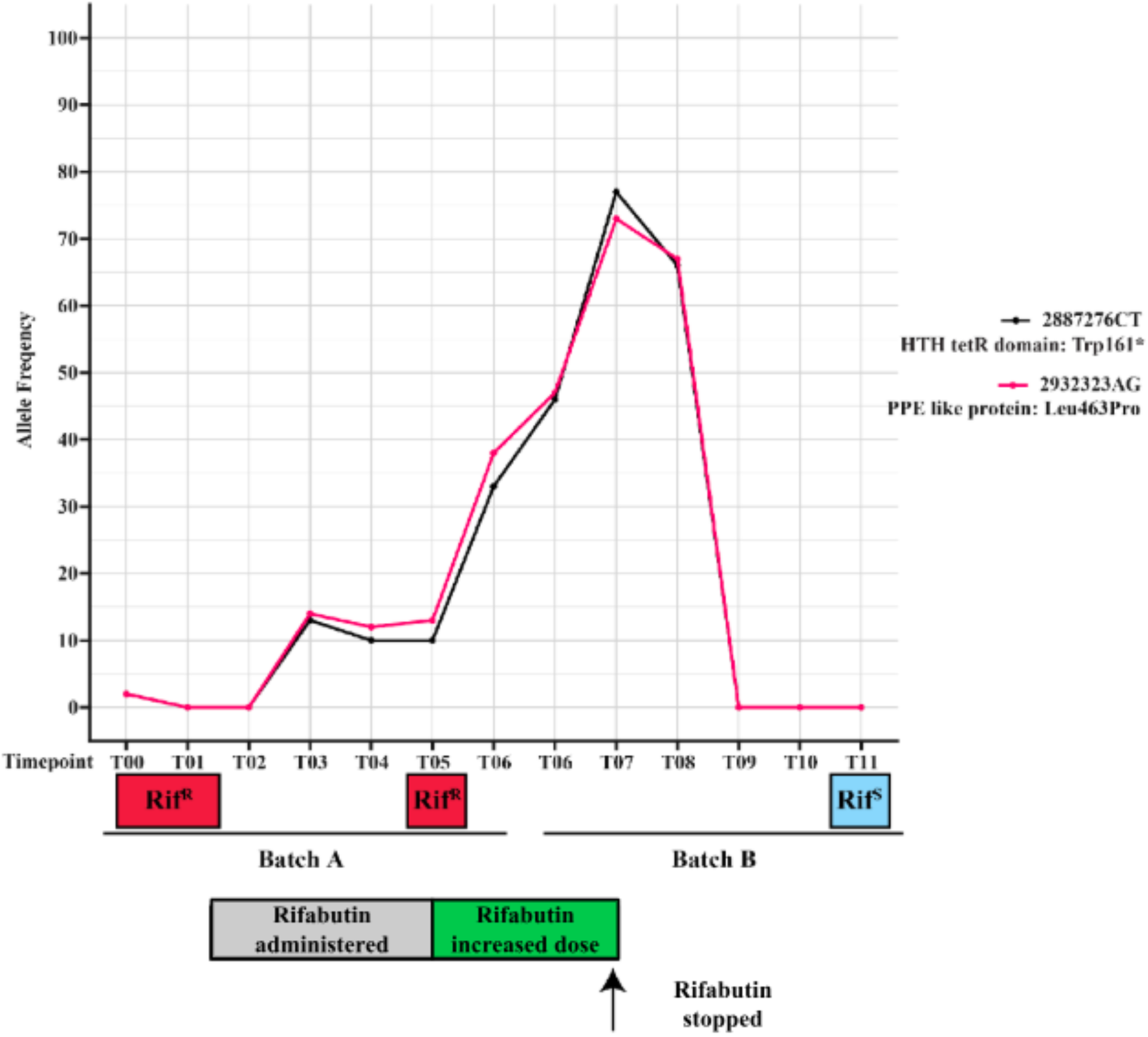
Dynamics of SNPs 2887276CT and 2932323AG as potential rifampicin resistance mutations. Information on drug susceptibility to relevant antibiotics is provided below, with the light blue representing susceptibility to a drug and red representing resistance. Rifabutin treatment was started between T01 and T02 (grey box), increasing doses between T05 and T07 (green box), stopping around T07.

## References

1. Henkle E, Winthrop KL. Nontuberculous mycobacteria infections in immunosuppressed hosts. Clin Chest Med. 2015;36(1):91–9.

2. Ong LT. Epidemiology of nontuberculous mycobacteria infection in Asia: A narrative review. Indian J Tuberc. 2025;72(2):259–65.

3. Henkle E, Hedberg K, Schafer S, Novosad S, Winthrop KL. Population-based Incidence of Pulmonary Nontuberculous Mycobacterial Disease in Oregon 2007 to 2012. Ann Am Thorac Soc. 2015;12(5):642–7.

4. Jamal F, Hammer MM. Nontuberculous Mycobacterial Infections. Radiol Clin North Am. 2022;60(3):399–408.

5. Nguyen M-VH, Haas MK, Kasperbauer SH, Calado Nogueira de Moura V, Eddy JJ, Mitchell JD, et al. Nontuberculous Mycobacterial Pulmonary Disease: Patients, Principles, and Prospects. Clinical Infectious Diseases. 2024;79(4):e27–e47.

6. Prevots DR, Shaw PA, Strickland D, Jackson LA, Raebel MA, Blosky MA, et al. Nontuberculous mycobacterial lung disease prevalence at four integrated health care delivery systems. Am J Respir Crit Care Med. 2010;182(7):970–6.

7. Dahl VN, Mølhave M, Fløe A, van Ingen J, Schön T, Lillebaek T, et al. Global trends of pulmonary infections with nontuberculous mycobacteria: a systematic review. Int J Infect Dis. 2022;125:120–31.

8. Abate G, Stapleton JT, Rouphael N, Creech B, Stout JE, El Sahly HM, et al. Variability in the Management of Adults With Pulmonary Nontuberculous Mycobacterial Disease. Clin Infect Dis. 2021;72(7):1127–37.

9. Im Y, Choe J, Kim DH, Kim S-Y, Jhun BW. Treatment Outcomes of Mycobacterium avium Complex Pulmonary Disease with a 2-drug Daily Regimen Using Macrolide and Ethambutol. Open Forum Infectious Diseases. 2025;12(6).

10. Abate G, Stapleton JT, Rouphael N, Creech B, Stout JE, El Sahly HM, et al. Variability in the Management of Adults With Pulmonary Nontuberculous Mycobacterial Disease. Clinical Infectious Diseases. 2020;72(7):1127–37.

11. Adjemian J, Prevots DR, Gallagher J, Heap K, Gupta R, Griffith D. Lack of Adherence to Evidence-based Treatment Guidelines for Nontuberculous Mycobacterial Lung Disease. Annals of the American Thoracic Society. 2014;11(1):9–16.

12. Cassidy PM, Hedberg K, Saulson A, McNelly E, Winthrop KL. Nontuberculous mycobacterial disease prevalence and risk factors: a changing epidemiology. Clin Infect Dis. 2009;49(12):e124–9.

13. Chai J, Zhang S, Ma C, Mei Q, Liu T, Liu J, et al. Clinical analysis and risk factors associated with poor prognosis in nontuberculous mycobacterial infection. Virulence. 2025;16(1):2459313.

14. Erasmus JJ, McAdams HP, Farrell MA, Patz EF. Pulmonary Nontuberculous Mycobacterial Infection: Radiologic Manifestations. RadioGraphics. 1999;19(6):1487–503.

15. Guglielmi VE, Cummings JE, Whittel NJ, Langland EA, Slayden RA. NTM-host matched infection models for the classification of drug efficacy against rapid and slow growing nontuberculous mycobacteria species. Sci Rep. 2026;16(1).

16. García-Pedrazuela M, Frutos JM, Muñoz-Egea MC, Alcaide F, Tórtola T, Vitoria A, et al. Polyclonality among clinical strains of non-pigmented rapidly growing mycobacteria: phenotypic and genotypic differences and their potential implications. Clin Microbiol Infect. 2015;21(4):348.e1–4.

17. Oliveira RS, Sircili MP, Ueki SY, Telles MA, Schnabel B, Briones MR, et al. PCR-restriction enzyme analysis of a bone marrow isolate from a human immunodeficiency virus-positive patient discloses polyclonal infection with two Mycobacterium avium strains. J Clin Microbiol. 2000;38(12):4643–5.

18. Wallace RJ, Jr., Zhang Y, Brown BA, Dawson D, Murphy DT, Wilson R, et al. Polyclonal Mycobacterium avium complex infections in patients with nodular bronchiectasis. Am J Respir Crit Care Med. 1998;158(4):1235–44.

19. van Ingen J, Egelund EF, Levin A, Totten SE, Boeree MJ, Mouton JW, et al. The pharmacokinetics and pharmacodynamics of pulmonary Mycobacterium avium complex disease treatment. Am J Respir Crit Care Med. 2012;186(6):559–65.

20. Ernest JP, Sarathy J, Wang N, Kaya F, Zimmerman MD, Strydom N, et al. Lesion Penetration and Activity Limit the Utility of Second-Line Injectable Agents in Pulmonary Tuberculosis. Antimicrob Agents Chemother. 2021;65(10):e0050621.

21. Rodriguez-Pazmiño AS, Carvajal E, Paredes-Núñez D, Echeverría J, Calderon J, Orlando SA, et al. Comparative evaluation of MALDI-ToF mass spectrometry and Sanger sequencing of the 16S, hsp65, and rpoB genes for non tuberculous mycobacteria species identification. Front Cell Infect Microbiol. 2025;15:1612459.

22. Iwen PC, Tarantolo SR, Mohamed AM, Hinrichs SH. First report of Mycobacterium nebraskense as a cause of human infection. Diagn Microbiol Infect Dis. 2006;56(4):451–3.

23. Metersky ML, Losier AJ, Fraulino DA, Warnock TA, Varley CD, Le AM, et al. Mycobacterium nebraskense Isolated from Patients in Connecticut and Oregon, USA. Emerg Infect Dis. 2025;31(3):507–15.

24. Walsh AM, Roycroft E, Hinchion K, Basdeo SA, Sheedy FJ, Crispie F, et al. Genomic characterisation of recurrent Mycobacterium avium isolates from chronically infected patients reveals patterns of within-host evolution. Genome Medicine. 2025;17(1):120.

25. Meier A, Kirschner P, Springer B, Steingrube VA, Brown BA, Wallace RJ, Jr., et al. Identification of mutations in 23S rRNA gene of clarithromycin-resistant Mycobacterium intracellulare. Antimicrob Agents Chemother. 1994;38(2):381–4.

26. Moazed D, Noller HF. Chloramphenicol, erythromycin, carbomycin and vernamycin B protect overlapping sites in the peptidyl transferase region of 23S ribosomal RNA. Biochimie. 1987;69(8):879–84.

27. Versalovic J, Shortridge D, Kibler K, Griffy MV, Beyer J, Flamm RK, et al. Mutations in 23S rRNA are associated with clarithromycin resistance in Helicobacter pylori. Antimicrobial Agents and Chemotherapy. 1996;40(2):477–80.

28. Vester B, Douthwaite S. Macrolide Resistance Conferred by Base Substitutions in 23S rRNA. Antimicrobial Agents and Chemotherapy. 2001;45(1):1–12.

29. Skinner R, Cundliffe E, Schmidt FJ. Site of action of a ribosomal RNA methylase responsible for resistance to erythromycin and other antibiotics. J Biol Chem. 1983;258(20):12702–6.

30. Menninger JR. Functional consequences of binding macrolides to ribosomes. J Antimicrob Chemother. 1985;16 Suppl A:23–34.

31. Walker TM, Miotto P, Köser CU, Fowler PW, Knaggs J, Iqbal Z, et al. The 2021 WHO catalogue of Mycobacterium tuberculosis complex mutations associated with drug resistance: A genotypic analysis. Lancet Microbe. 2022;3(4):e265–e73.

32. Chien JY, Chiu WY, Chien ST, Chiang CJ, Yu CJ, Hsueh PR. Mutations in gyrA and gyrB among Fluoroquinolone- and Multidrug-Resistant Mycobacterium tuberculosis Isolates. Antimicrob Agents Chemother. 2016;60(4):2090–6.

33. Kocagöz T, Hackbarth CJ, Unsal I, Rosenberg EY, Nikaido H, Chambers HF. Gyrase mutations in laboratory-selected, fluoroquinolone-resistant mutants of Mycobacterium tuberculosis H37Ra. Antimicrob Agents Chemother. 1996;40(8):1768–74.

34. Mayer C, Takiff H. The Molecular Genetics of Fluoroquinolone Resistance in *Mycobacterium tuberculosis*. Microbiology Spectrum. 2014;2(4):10.1128/microbiolspec.mgm2-0009-2013.

35. Stamm LV, Greene SR, Barnes NY. Identification and characterization of the gyrB gene from Treponema pallidum subsp. pallidum. FEMS Microbiol Lett. 1997;153(1):129–34.

36. Organization WH. Catalogue of mutations in Mycobacterium tuberculosis complex and their association with drug resistance: World Health Organization; 2023.

37. Guiraud J, Piau C, Enault C, Nkpa Charron E, Ducos D, Lafuente C, et al. Comparison of the molecular FluoroType Mycobacteria VER 1.0 and the Maldi BioTyper Mycobacteria assays for the identification of non-tuberculous mycobacteria. J Clin Microbiol. 2025;63(1):e0120624.

38. Moon SM, Park HY, Kim SY, Jhun BW, Lee H, Jeon K, et al. Clinical Characteristics, Treatment Outcomes, and Resistance Mutations Associated with Macrolide-Resistant Mycobacterium avium Complex Lung Disease. Antimicrob Agents Chemother. 2016;60(11):6758–65.

39. Matzneller P, Krasniqi S, Kinzig M, Sörgel F, Hüttner S, Lackner E, et al. Blood, tissue, and intracellular concentrations of azithromycin during and after end of therapy. Antimicrob Agents Chemother. 2013;57(4):1736–42.

40. Christianson S, Grierson W, Wolfe J, Sharma MK. Rapid molecular detection of macrolide resistance in the Mycobacterium avium complex: are we there yet? J Clin Microbiol. 2013;51(7):2425–6.

41. Alffenaar JW, Märtson AG, Heysell SK, Cho JG, Patanwala A, Burch G, et al. Therapeutic Drug Monitoring in Non-Tuberculosis Mycobacteria Infections. Clin Pharmacokinet. 2021;60(6):711–25.

42. Daley CL, Iaccarino JM, Lange C, Cambau E, Wallace RJ, Jr., Andrejak C, et al. Treatment of nontuberculous mycobacterial pulmonary disease: an official ATS/ERS/ESCMID/IDSA clinical practice guideline. Eur Respir J. 2020;56(1).

43. Du X, Shi K, Zhang H, Chong Y. Screening for heterogeneous drug resistance in tuberculosis and its impact on clinical prognosis: A comprehensive review. iScience. 2026;29(3):115049.

44. Haworth CS, Banks J, Capstick T, Fisher AJ, Gorsuch T, Laurenson IF, et al. British Thoracic Society Guideline for the management of non-tuberculous mycobacterial pulmonary disease (NTM-PD). BMJ Open Respir Res. 2017;4(1):e000242.

45. Hong SK, Kim EC. Possible misidentification of Mycobacterium yongonense. Emerg Infect Dis. 2014;20(6):1089–90.

46. Mougari F, Loiseau J, Veziris N, Bernard C, Bercot B, Sougakoff W, et al. Evaluation of the new GenoType NTM-DR kit for the molecular detection of antimicrobial resistance in non-tuberculous mycobacteria. J Antimicrob Chemother. 2017;72(6):1669–77.

47. Woods GL, Clinical,. LSI. Susceptibility Testing of Mycobacteria, Nocardiae, and Other Aerobic Actinomycetes: Approved Standard: Clinical and Laboratory Standards Institute; 2011.

48. Arefin MS, Mitu MJ, Mitu SY, Nurjahan A, Mobin M, Nahar S, et al. Mutational alterations in the QRDR regions associated with fluoroquinolone resistance in Pseudomonas aeruginosa of clinical origin from Savar, Dhaka. PLoS One. 2025;20(2):e0302352.

49. Deguchi T, Yasuda M, Nakano M, Ozeki S, Ezaki T, Maeda S, et al. Rapid detection of point mutations of the Neisseria gonorrhoeae gyrA gene associated with decreased susceptibilities to quinolones. J Clin Microbiol. 1996;34(9):2255–8.

50. Dessus-Babus S, Bébéar Cécile M, Charron A, Bébéar C, de Barbeyrac B. Sequencing of Gyrase and Topoisomerase IV Quinolone-Resistance-Determining Regions of Chlamydia trachomatis and Characterization of Quinolone-Resistant Mutants Obtained In Vitro. Antimicrobial Agents and Chemotherapy. 1998;42(10):2474–81.

51. Devasia R, Blackman A, Eden S, Li H, Maruri F, Shintani A, et al. High Proportion of Fluoroquinolone-Resistant Mycobacterium tuberculosis Isolates with Novel Gyrase Polymorphisms and a gyrA Region Associated with Fluoroquinolone Susceptibility. Journal of Clinical Microbiology. 2012;50(4):1390–6.

52. Hooper DC, Jacoby GA. Topoisomerase Inhibitors: Fluoroquinolone Mechanisms of Action and Resistance. Cold Spring Harb Perspect Med. 2016;6(9).

53. Cha SB, Jeon BY, Kim WS, Kim JS, Kim HM, Kwon KW, et al. Experimental Reactivation of Pulmonary Mycobacterium avium Complex Infection in a Modified Cornell-Like Murine Model. PLoS One. 2015;10(9):e0139251.

54. Maslow JN, Brar I, Smith G, Newman GW, Mehta R, Thornton C, et al. Latent infection as a source of disseminated disease caused by organisms of the Mycobacterium avium complex in simian immunodeficiency virus-infected rhesus macaques. J Infect Dis. 2003;187(11):1748–55.

55. Clinic M. <Culture_Referred_for_Identification_and_Susceptibility_for_Mycobacterium_and_Nocardia_A lgorithm.pdf>.

56. Galveston UT. <lab-testing-procedures-05-31-24.pdf>. 2024.

57. Beyvers S, Jelonek L, Goesmann A, Schwengers O. Bakta Web – rapid and standardized genome annotation on scalable infrastructures. Nucleic Acids Research. 2025;53(W1):W51–W6.

58. Schwengers O, Jelonek L, Dieckmann MA, Beyvers S, Blom J, Goesmann A. Bakta: rapid and standardized annotation of bacterial genomes via alignment-free sequence identification. Microbial Genomics. 2021;7(11).

59. Hunt M, Silva ND, Otto TD, Parkhill J, Keane JA, Harris SR. Circlator: automated circularization of genome assemblies using long sequencing reads. Genome Biol. 2015;16:294.

60. Huerta-Cepas J, Szklarczyk D, Heller D, Hernández-Plaza A, Forslund SK, Cook H, et al. eggNOG 5.0: a hierarchical, functionally and phylogenetically annotated orthology resource based on 5090 organisms and 2502 viruses. Nucleic Acids Res. 2019;47(D1):D309–d14.

61. Ciufo S, Kannan S, Sharma S, Badretdin A, Clark K, Turner S, et al. Using average nucleotide identity to improve taxonomic assignments in prokaryotic genomes at the NCBI. Int J Syst Evol Microbiol. 2018;68(7):2386–92.

62. Konstantinidis KT, Tiedje JM. Genomic insights that advance the species definition for prokaryotes. Proc Natl Acad Sci U S A. 2005;102(7):2567–72.

63. Wishart DS, Han S, Saha S, Oler E, Peters H, Grant Jason R, et al. PHASTEST: faster than PHASTER, better than PHAST. Nucleic Acids Research. 2023;51(W1):W443–W50.

64. Ondov BD, Treangen TJ, Melsted P, Mallonee AB, Bergman NH, Koren S, et al. Mash: fast genome and metagenome distance estimation using MinHash. Genome Biology. 2016;17(1):132.

65. Chen S. Ultrafast one-pass FASTQ data preprocessing, quality control, and deduplication using fastp. iMeta. 2023;2(2):e107.

66. Okonechnikov K, Conesa A, García-Alcalde F. Qualimap 2: advanced multi-sample quality control for high-throughput sequencing data. Bioinformatics. 2016;32(2):292–4.

67. Benjamin D, Sato T, Cibulskis K, Getz G, Stewart C, Lichtenstein L. Calling Somatic SNVs and Indels with Mutect2. bioRxiv. 2019:861054.

68. Wilm A, Aw PP, Bertrand D, Yeo GH, Ong SH, Wong CH, et al. LoFreq: a sequence-quality aware, ultra-sensitive variant caller for uncovering cell-population heterogeneity from high-throughput sequencing datasets. Nucleic Acids Res. 2012;40(22):11189–201.

69. Koboldt DC, Zhang Q, Larson DE, Shen D, McLellan MD, Lin L, et al. VarScan 2: somatic mutation and copy number alteration discovery in cancer by exome sequencing. Genome Res. 2012;22(3):568–76.

70. Foundation PS. Python. Python Software Foundation; 2026.

71. Ikotun AM, Ezugwu AE, Abualigah L, Abuhaija B, Heming J. K-means clustering algorithms: A comprehensive review, variants analysis, and advances in the era of big data. Information Sciences. 2023;622:178–210.

72. Kassambara AaM, Fabian. factoextra: Extract and Visualize the Results of Multivariate Data Analyses. In: With contributions from Laszlo Erdey (Faculty of Economics and Business UoD, Hungary), editor.: R package version 2.0.0.; 2026.

73. Deitrick C. Lolipop manual. Github; 2020.

74. Lieberman TD, Flett KB, Yelin I, Martin TR, McAdam AJ, Priebe GP, et al. Genetic variation of a bacterial pathogen within individuals with cystic fibrosis provides a record of selective pressures. Nat Genet. 2014;46(1):82–7.

75. Klug WS, Cummings MR, Spencer CA, Palladino MA, Killian D. Essentials of Genetics. 10 ed: Pearson Education; 2020.

